# Periodic variations in the Covid-19 infection and fatality rates

**DOI:** 10.1101/2020.09.08.20190751

**Authors:** S. Peirani, Ph. Icard, J.A. de Freitas Pacheco

## Abstract

Power spectra of infection and mortality rate curves for nineteen countries of different continents were computed. Nine of them show the presence of oscillations with a period of about seven says either in the infection or in the mortality data sets. The computed power spectra for seven countries do no indicate any significant signal of periodicity while the three remaining countries indicate periodic oscillations only in the infection or only in the mortality curve. Data indicate that minima occur generally on weekends. The seven-day periodicity present in infection data of nine countries is robust and seems to be the consequence of different factors as, for instance, higher testing frequency during weekdays or/and an enhanced contamination during social activities during weekends. For the moment, there is no convincing explanation for the seven-day oscillations observed in the mortality curves of some countries.

## 1. Introduction

Presently almost all countries in the world are affected by the SARS-CoV-2 epidemic. Up to now (2020–08–27), according to the World Health Organization (WHO), about 24 millions of persons have been contaminated and approximately 821 000 casualties have been notified.

The absence of a vaccine or of an efficient treatment for this disease led to health officials of different countries to adopt specific strategies to control the propagation of the virus. Many countries, in particular in Europe, inspired by the success obtained by the Chinese authorities in the control of the virus spread in Wuhan, adopted a lockdown procedure aiming to reduce the so-called R_0_ parameter, i.e., the average number of infected persons per a single sick individual.

In countries in which the majority of the population followed the adopted lockdown rules, the initial exponential growth of the number of cases was controlled. In those countries, after attaining a maximum, a slow decrease either in the infection or in the fatality rate was observed. This behavior is clearly seen, for instance, in the evolution of the pandemic in China, France, Spain, Italy, Germany, Canada, among others. However, at the present time the number of cases is still growing in countries like India, Iraq, USA, Brazil, among others. In some countries the evolution does not follow the “standard” picture once the curve describing the infection rate deviates considerably from the well-known “bell-shape”. The Swedish have not adopted a lockdown procedure, deciding to control the epidemic based on a “collective immunity” but the health authorities searched to protect fragile people like seniors or persons having risk factors like diabetes and obesity. The infection rate evolution in Sweden had an initial exponential growth until end of April and then stabilized at a rate of about 600 new cases per day. However, at the end of May, the number of cases increased again up to 1100 infection per day, indicating that the adopted strategy of “collective immunity” is disputable. It is worth mentioning that this “raising” effect seen in the number of new cases has not affected significantly the fatality rate, indicating that the population being presently contaminated is constituted by “young” individuals in its majority. The same situation is presently also occurring in France.

However a particular feature seems to be present either in the infection or in the fatality rate evolutionary curves of some countries, i.e., oscillations are superimposed on the average rates. In fact, a recent study [1] based on autocorrelation data analysis has shown that these oscillations are periodic with a period of about seven days. The authors of such an investigation interpreted their result as a consequence of inter-generational social interactions occurring during the weekend. In these social events, a transfer of the virus from asymptomatic young to old vulnerable individuals would occur. According to [1], these regular contamination episodes would be responsible for the periodic structures observed in the new daily cases curve. A more robust mathematical technique was adopted by [2] for the search of periodicities in data of USA as well as in localized regions like the cities of New York and Los Angeles. In this study, the density power spectrum (DPS) of time series was computed for the aforementioned data and the authors found that variations present in the daily new cases curve of the two American cities are correlated with the frequency of testing. In other words, during the weekdays more tests are performed than during the weekends and this effect would be responsible for the observed oscillations.

Here we report our search for periodicities in the infection and mortality data of nineteen countries distributed around different continents by using the DPS technique. We found a significant seven-day signal in nine countries (USA, Brazil, France, Mexico, UK, Italy, Germany, Spain and South Africa) while seven other countries (Algeria, Morocco, Tunisia, India, Egypt, Turkey and Iran) do not show any periodic signal in their DPS. Two countries (Belgium and Sweden) show the seven-day signal in the infection curve but any periodic oscillation is seen in the fatality curves. Russia was the only country to present a significant seven-day signal in the mortality data that is absent in the infection rate curve. In the next section the characteristics of the aforementioned variations will be discussed as well as possible explanations for them.

## 2. Periodic variations in infection and mortality rates

As mentioned above, seven-day oscillations are present either in the infection or in the fatality rate curves of nine countries among the nineteen included in our study. In order to illustrate these oscillations,figure 1 shows the evolution of the infection and fatality rates observed in France, Brazil, Italy and USA (data taken respectively from refs. [3], [4], [5] and [6]. When data references were not explicitly mentioned, they were taken from “https://www.worldometer.info/coronavirus/”). Notice that infection rates were divided by a factor of ten in order to plot both curves in the same scale.

**Figure 1.**
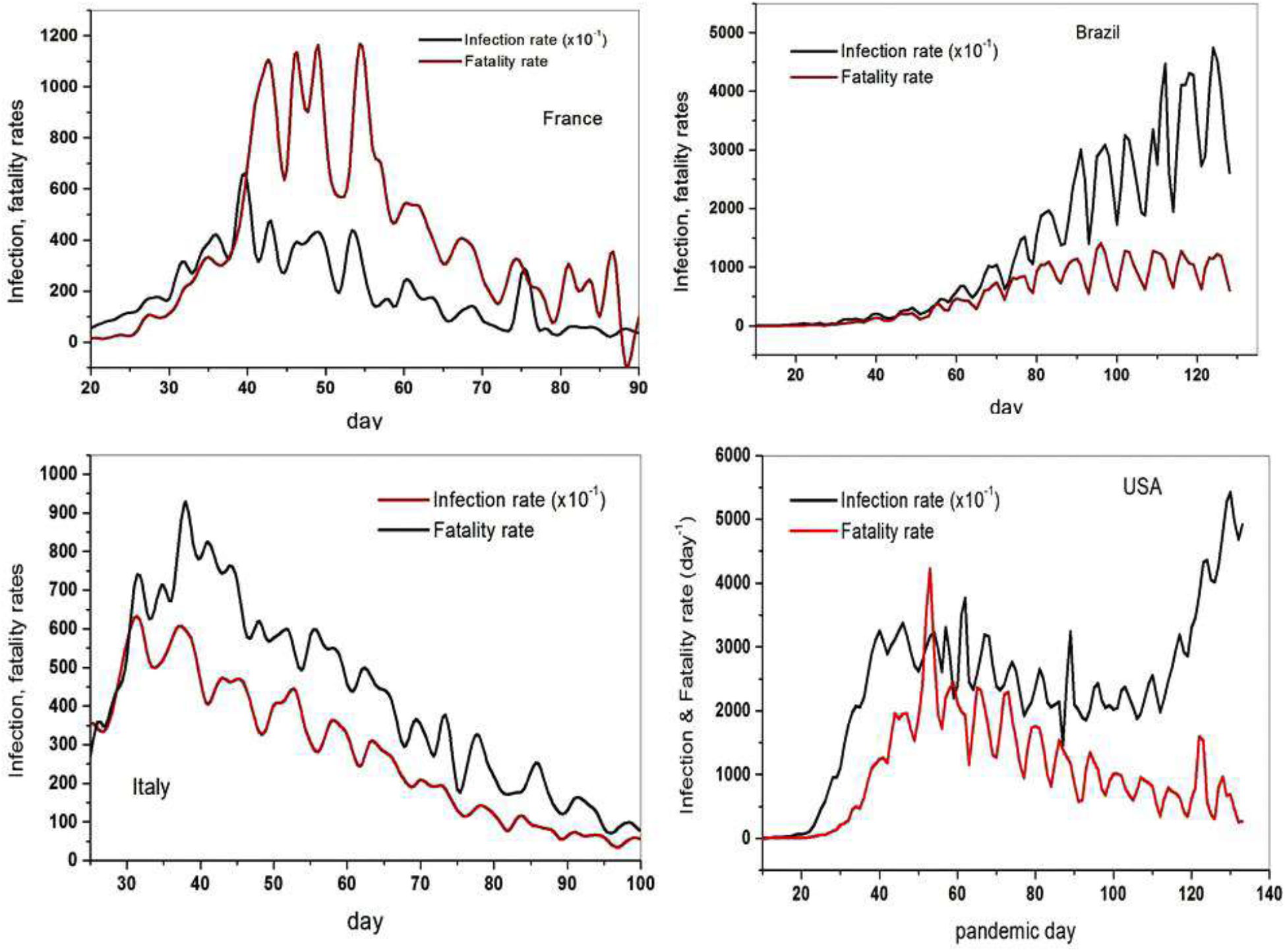
Evolution of infection and fatality rates for France, Brazil, Italy and USA. Notice that to obtain the true infection rate, values must be multiplied by a factor of 10 and that minima of both curves are generally coincident.

Simple inspection of figure 1 indicates that minima present in the evolution of both infection and fatality rates practically coincide in the case of France, Brazil and USA but sometimes a small phase lag is observed in data from Italy. Minima or valleys occur generally during the weekend in all these countries as already noticed by [1]. In this case, the explanation proposed by [2], i.e., tests to detect contaminated individuals are more often made on weekdays than on weekends could be a possible explanation for the presence of such oscillations in the contamination curves.

**Figure 2.**
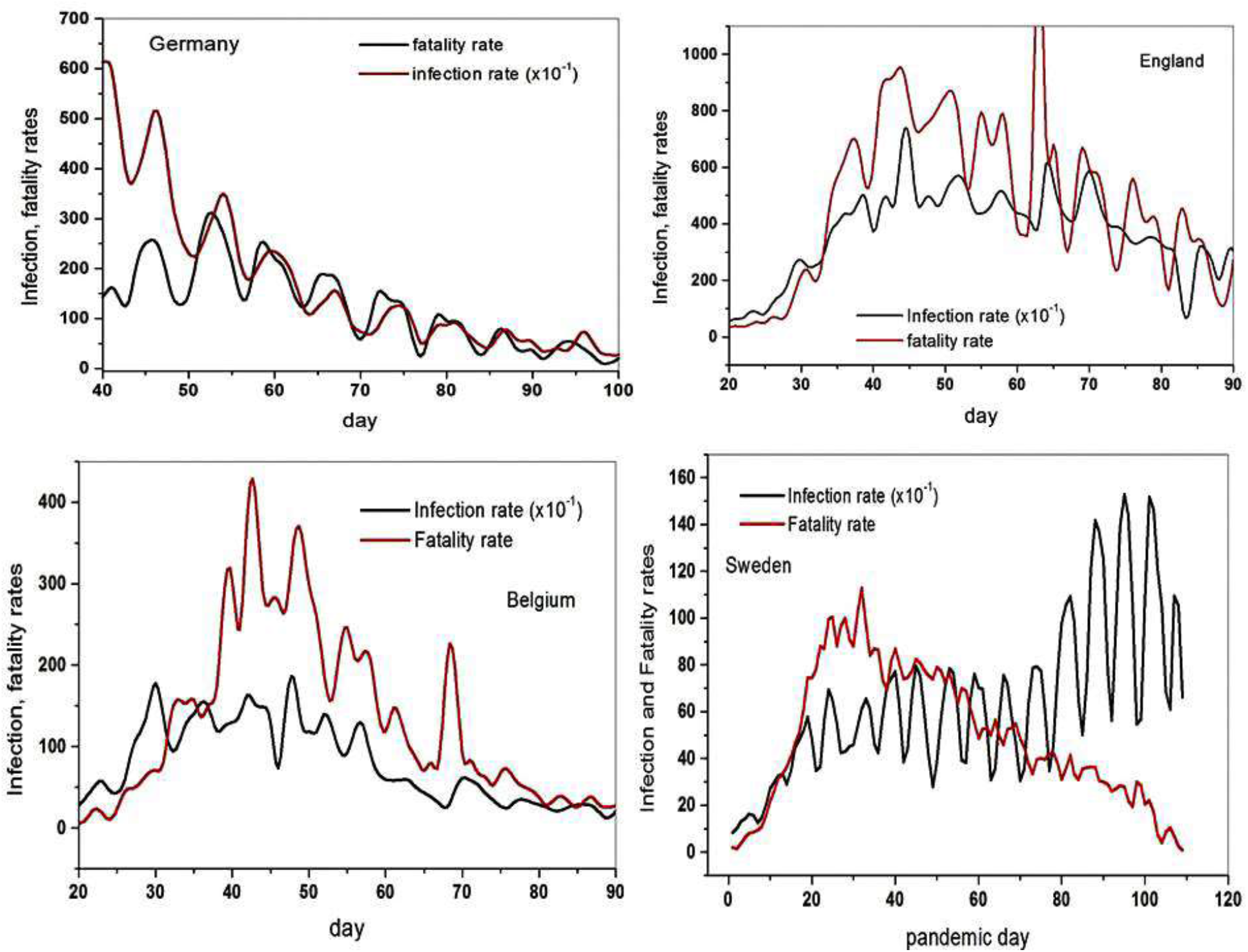
Evolution of the infection and fatality rates for Germany, England, Belgium and Sweden. Note the different behavior with respect to the curves displayed in the panels of figure 1.

Figure 2 shows the same curves as in figure 1 but for Germany, England, Belgium and Sweden. We notice first that for Germany and England the valleys observed in both curves are in general not coincident, having a lag of one or even two days in different epidemic epochs. Consequently, as we shall see, the oscillation periods derived from the DPS analysis of the infection and fatality curves are not rigorously the same. For Belgium the correlation is very poor since oscillations in the mortality rate are barely noticeable. In the case of Sweden, despite the modulation be clearly seen in the infection rate it is practically undetectable in the fatality rate curve. As mentioned before, the different behavior of the two curves after day 80 in the Swedish data suggests that probably a young population is being contaminated, which causes the increasing number of daily new cases while the fatality rate curve is reflecting essentially what is occurring among seniors and fragile individuals. Such a population difference could be a possible explanation for the weak correlation observed between both curves in Swedish data.

In order to check the significance of these oscillations and to have a better estimation of their period, we have computed the DPS for the available data of those nineteen countries by using a Fast Fourier Transform (FFT) technique (see, for instance, [7]). Countries showing oscillations in the infection and fatality rates nearly in phase (minimum occurring at weekends) the DPS presents a well-defined pic (sometimes also the first harmonic) whose period derived either from the infection or the mortality curve is practically the same.

**Figure 3.**
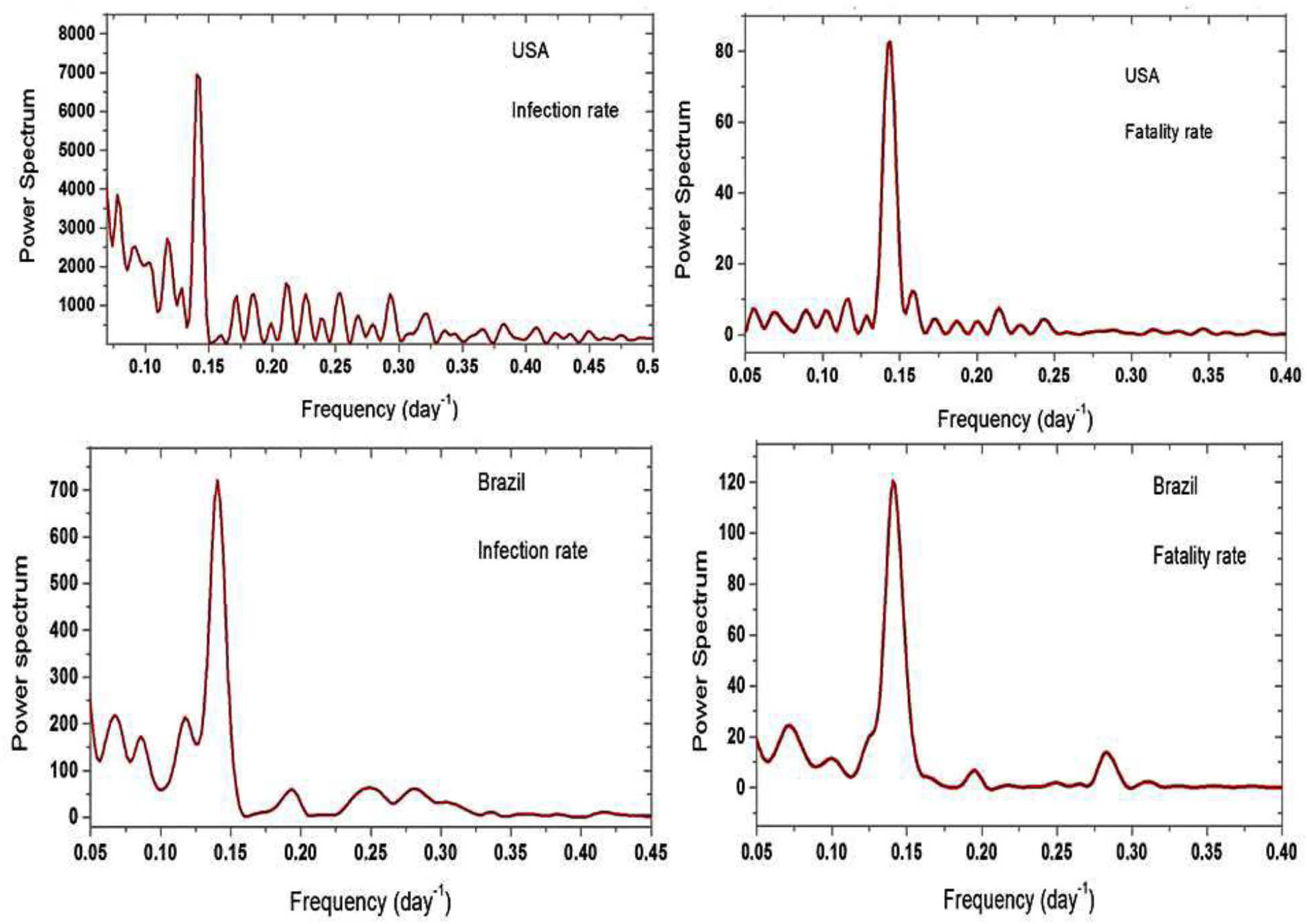
Density power spectra derived for daily new cases and fatality data available from USA and Brazil.

Table 1 summarizes the results for the nine countries where the seven days signal has been found in both infection and mortality data sets. Again, to give examples of the computed DPS, figure 3 shows the resulting power spectra for USA and Brazil. For USA, the main pic is at 7.04 days for the infection and 6.97 days for the fatality data while for Brazil the main pics are observed at 7.10 and 7.09 days respectively for infection and fatality rates. Notice that the pics have amplitudes well above the noise. These results are consistent with the concordance of the minimum instants observed in figure 1.

**Table 1.**
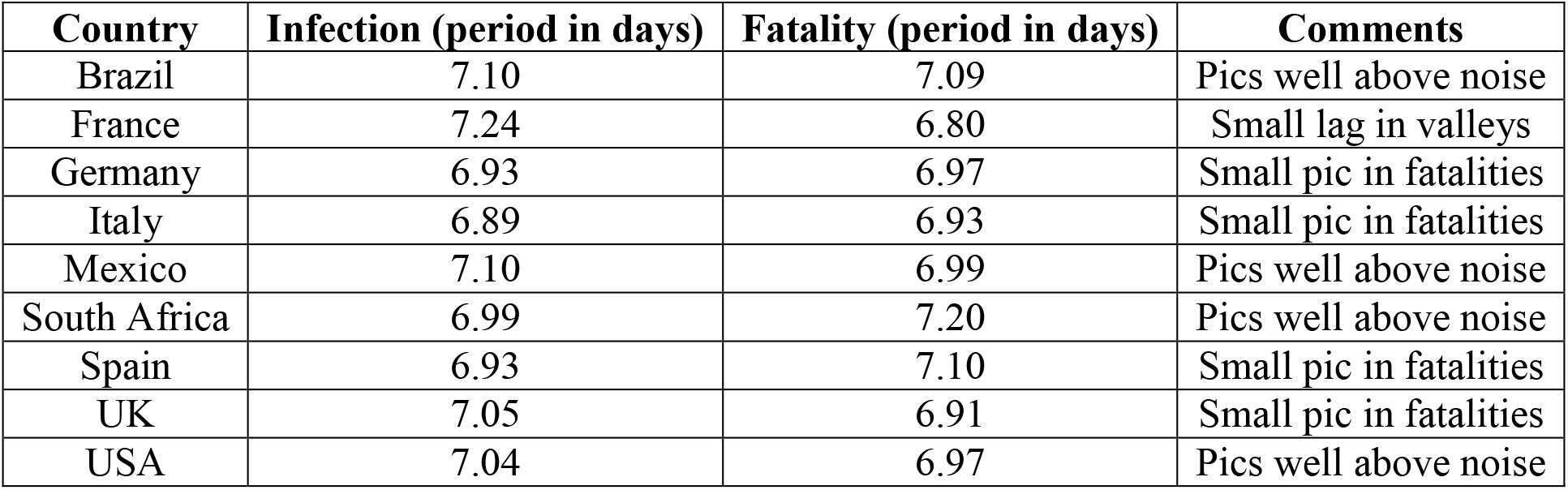
The first column gives the country, the following two columns give the periods (in days) derived from the DPS respectively from the infection and fatality rates data. Finally, the last column gives some comments on the relative amplitude of the pics.

| Country | Infection (period in days) | Fatality (period in days) | Comments |
| --- | --- | --- | --- |
| Brazil | 7.10 | 7.09 | Pics well above noise |
| France | 7.24 | 6.80 | Small lag in valleys |
| Germany | 6.93 | 6.97 | Small pic in fatalities |
| Italy | 6.89 | 6.93 | Small pic in fatalities |
| Mexico | 7.10 | 6.99 | Pics well above noise |
| South Africa | 6.99 | 7.20 | Pics well above noise |
| Spain | 6.93 | 7.10 | Small pic in fatalities |
| UK | 7.05 | 6.91 | Small pic in fatalities |
| USA | 7.04 | 6.97 | Pics well above noise |

**Figure 4.**
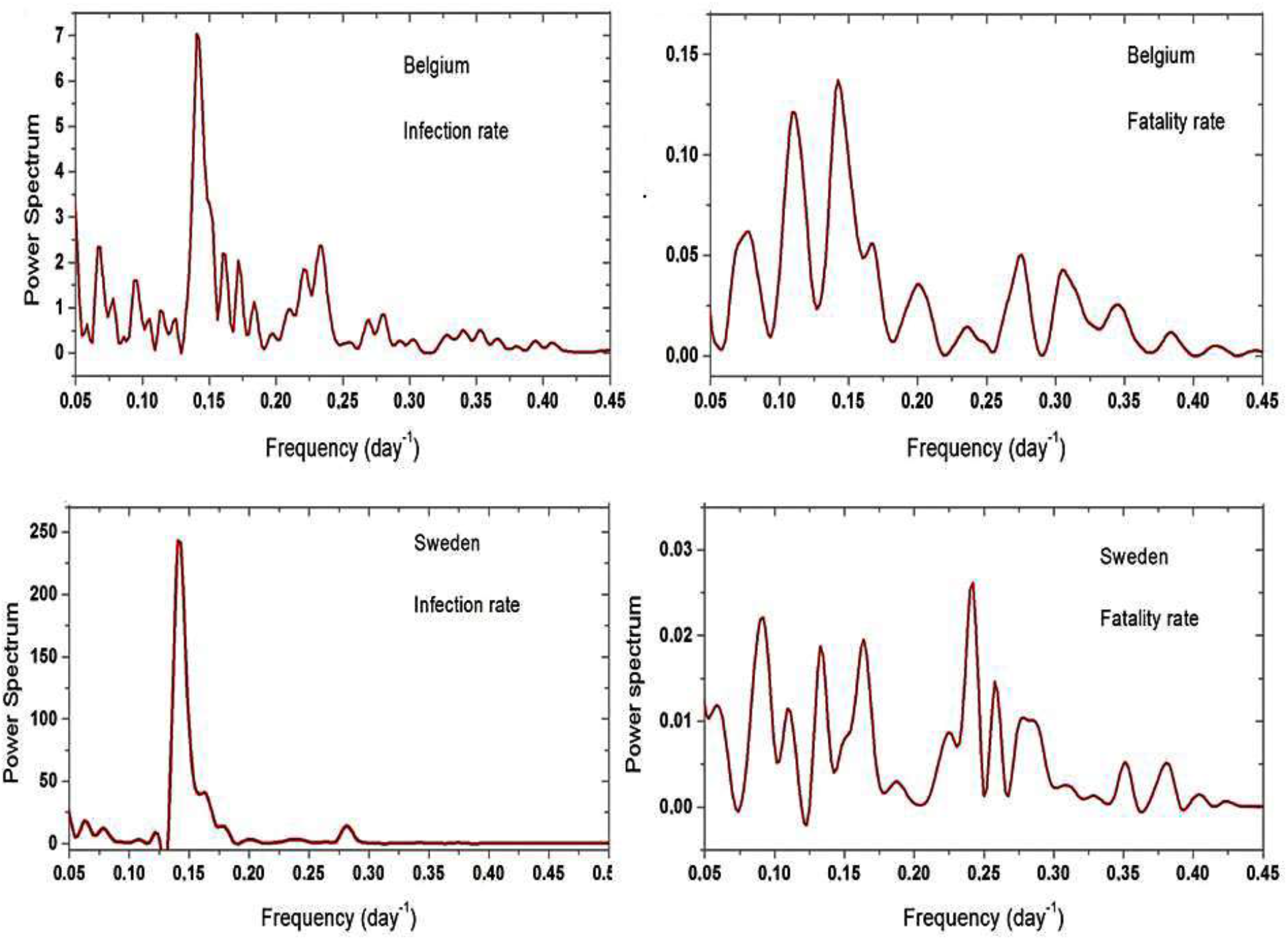
Density power spectra derived from infection and fatality rates for Belgium and Sweden

In figure 4 are shown the DPS derived for two countries (Belgium and Sweden) for which the signal is clear only for the infection rate curve. Notice that for these two countries the apparent minima observed in infection and fatality curves do not coincide. The pics are significantly well above the noise (in particular for Sweden) when the infection data are considered but this is not the case for the fatality curves. The DPS for Belgium indicates a period of 7.06 days while for Sweden data the derived period is 7.09 days.

In order to compare our numerical procedure (FFT) with the computations reported by [2], we have evaluated the DPS for the New York City data set. Our derived DPSs confirm the previous results by [2], that is, oscillations with a period of 6.96 days are present in the infection rate curve but not in that of the mortality rate. We emphasize that valleys in the NYC curve of daily cases occur also on weekends (mainly on Sundays).

**Figure 5.**
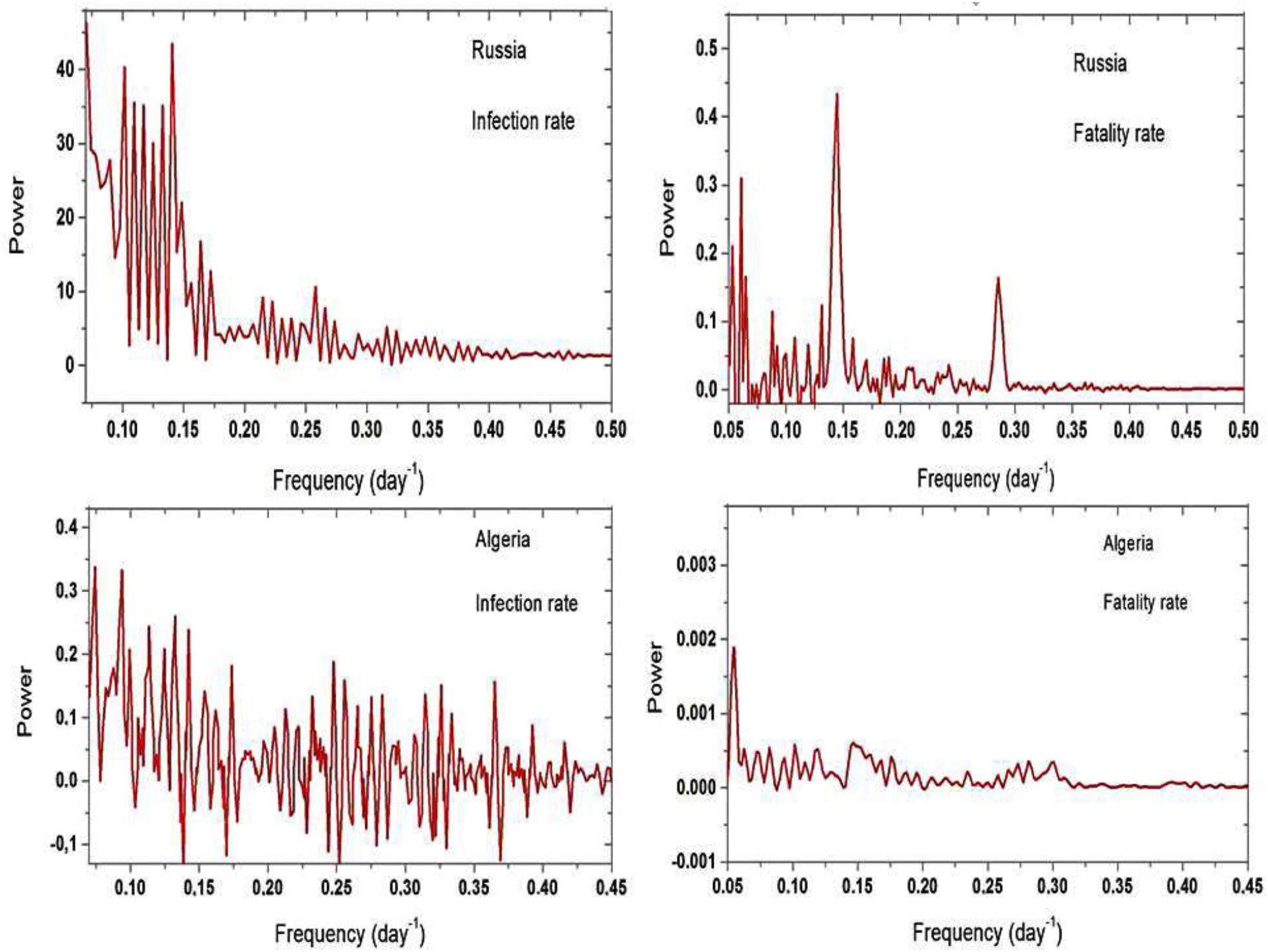
DPS for Russia (upper panels) and Algeria (lower panels). Notice a pure noise spectrum derived for the Algeria data and for the infection curve of Russia.

The DPS of seven other countries (Algeria, Morocco, Tunisia, India, Egypt, Turkey and Iran) do not show any periodic signal. Their DPS show a “pure” white noise spectrum. Russia is one exception, since it is the only country of our list for which a weak seven-day signal is present in the fatality curve but not in the infection evolution. To illustrate these last cases, the derived DPS for Algeria and Russia are shown in figure 5.

The oscillations appearing in data sets of different countries can be modeled by an effective modulated R_0_ parameter and we have tested such an approach by computing some epidemic propagation models based on Monte Carlo techniques. Although the details of these calculations are beyond the purposes of the present paper (they will be discussed elsewhere), for the sake of completeness we give here some basic guidelines of our numerical procedure.

For a given population, represented by N_0_ individuals, five distinct categories are identified: **A** – constituted by healthy individuals; **I** – infected but asymptomatic persons; **H** – infected persons with symptoms who are isolated at home or at a hospital; **R** – recovered individuals and **D** – deceased persons. In our scheme only individuals of class **I** can diffuse the virus and their contamination road is followed day by day during the incubation period. In a given day the number of persons contaminated by a single individual is decided by a Monte-Carlo procedure. The average number of contaminations (the R_0_ parameter) is supposed to decrease as the fraction of the healthy persons decreases in order to weaken the probability of contamination. The initial healthy population is assumed to have a given age distribution (in our simulations, the French age pyramid was adopted). Thus, each new contaminated person of a given age goes to class I and stays there during all incubation period, which may vary from three up to eleven days. Again, a Monte Carlo procedure decides the incubation period of each individual. After their incubation period, each individual is removed into class **H** where he can stay during an interval of 15–25 days. A Monte Carlo procedure determines the hospitalization time of each person and the probability of recovering or death is fixed by the age of the individual, which was taken from tables by **http://weekly.chinacdc.cn/en/article/id/e53946e2-c6c4-41e9-9a9b-fea8db1a8f51**. Thanks to this approach, it is possible to introduce a periodic modulation in the R_0_ parameter in order to simulate an enhanced contamination in a given day (or days) of the week. Such a modulation produces indeed a modulated infection rate evolution but it does not alter the rate of mortality. Hence, whatever is the reason (or reasons) producing the minima in the mortality rate curves, they should be distinct from those modulating the infection rate.

## 3. Final considerations

In the present investigation searches for periodic variations on data of infection and mortality rates of nineteen countries in different continents were performed, using the density power spectrum technique. We have detected the presence of a seven-day periodic signal in the data set of nine countries. Our findings are compatible with the results reported by [1] whose authors, by using a different approach, found these oscillations in data of seven countries among twelve investigated.

In the cases of Belgium, Germany, USA and UK both studies concluded that the seven-day signal in the mortality data. However, contrary to the claim by [1], our DPS analysis indicates that the signal is weak in the Swedish fatality data. However, it is worth mentioning that our investigation of the time series covers a longer interval than that used by [1] (up to April 28^th^ for ref.[1] and up to August 27^th^ in the present work). Nevertheless we point that both studies indicate that the seven-day oscillations are not a common feature present in data of all countries.

In general, the seven-day signal detected in the infection curves are more robust that that appearing sometimes in the mortality data. The investigation by [2] on the evolution of new cases in the cities of New York and Los Angeles suggests a significant correlation between variations in the rate of new cases with the frequency of testing. Since tests are preferentially made during weekdays, one should expect that minima would occur during weekends in the infection rate curve. This possibility fits with the absence of a seven-day signal in countries like Algeria, Morocco and Tunisia among others, where the daily number of tests realized is relatively small.

An additional possibility concerns the increased number of social activities during weekends, which combined with a median four-day incubation period (see [9]) leads to an increase of new cases during the middle of the week, consistent with the observed oscillation pattern. The case of India goes in this sense since a significant number of tests are being performed according to the health authorities but no seven-day signal appears in the infection curve. However, the lockdown rules are not strictly observed in India by the population, smoothing the presence of possible modulations in the infection rate evolution.

Probably a combination of different factors may contribute at different levels to the observed oscillations in the reported number of daily new cases. However, there is no convincing explanation for oscillations present in the mortality rates and the case of Russia is quite singular because the seven-day signal appears in the mortality data but not on the infection rate curve.

Our Monte Carlo simulations do not show any effect of a periodic modulation in the R_0_ parameters on the fatality rate evolution. The presence of seven-day oscillations in the mortality curve is less robust than that observed in the infection rate data. However, it is clear that the cause (or causes) responsible for oscillations in the daily infection data is (are) not expected to affect the mortality rate curve and distinct causes are necessary.

Some past investigations have analyzed the variations in medical care at hospitals between weekdays and weekends [8]. According to these authors, patients having procedures during weekends had higher mortality than those treated during weekdays. This possibility is certainly not responsible for the observed oscillations in the mortality rates because minima occur during weekends, contrary to the conclusions reached by [8]. It should be mentioned that in the case of Covid-19, a study on 191 patients in Wuhan (54 deceased) indicates that median time to death was 18.5 days [10]. Including an incubation period of about 4 days, the typical timescale between contamination and death is about 3 weeks and that could be a hint to explain the periodicity observed in the mortality curve of some countries.

Nevertheless, a suspicion was raised by [2] when comparing the mortality rate of the cities of New York and Los Angeles with that of USA as a whole. According to [2], the USA data set reflect the reported date of the casualties whereas the data set for New York and Los Angeles are back-dated to the casualty date. Consequently, the authors found (as we did) a significant seven days period on the mortality rate curve of the country but not in the data set of the two cities. The majority of the national health centers centralize the information received from local health headquarters and, in this case, the reported rates are simply dated as the information arrives, mainly in weekdays. According to [2], the oscillations in mortality rates would be probably an artifact caused by the way the information is collected by national centers. This possibility should be investigated more carefully by an analysis of the methods used by the national health agencies to collect and diffuse the statistical data.

## Data Availability

We used the evolution of the infection and fatality rates observed in different countries such as France, Brazil, Italy and USA for instance.

